# Vaccination exemptions among Kindergartners by state-level 2024 Presidential election result

**DOI:** 10.1101/2024.12.05.24318578

**Authors:** Jeremy Samuel Faust, Benjamin K. Renton

**Author notes:** **Corresponding author:** Jeremy Samuel Faust, MD, MS, Department of Emergency Medicine, Mass General Brigham, Division of Health Services Research, Harvard Medical School, Boston Massachusetts, 10 Vining St, Boston, MA 02115.

## Abstract

**Importance:** Pediatric vaccine exemptions have increased in recent years. How political environments influence these trends is unknown.

**Objective:** To analyze state-level kindergarten vaccination exemption rates from 2009-2024, stratified by 2024 Presidential election results, to assess changes over time.

**Design, Setting, and Participants:** This cross-sectional study used publicly available CDC data on kindergarten vaccination exemptions across all U.S. states and Washington, DC. States were categorized as Donald Trump or Kamala Harris states based on 2024 election results. Previous elections were also assessed.

**Main Outcomes and Measures:** Incident rates (IRs) of vaccination exemptions (all, medical, and non-medical) were calculated yearly. Incident rate ratios (IRRs) comparing vaccination exemption rates in Harris versus Trump win states were calculated. and Spearman correlations assessed associations between exemption rates and Republican vote share in the four most recent Presidential elections.

**Results:** Analysis included 53,997,748 person-years of kindergarten data. By 2023-2024, non-medical exemptions were permitted in 90.2% of states. Exemption rates increased in Trump states but decreased in Harris states during the study period. Most exemptions were non-medical.

**Conclusions and Relevance:** Kindergarten vaccination exemptions now diverge by political environment, increasing over time in states won by Trump in 2024 and decreasing in states won by Harris. The findings suggest growing politicization of vaccination practices, warranting targeted public health interventions.

## Introduction

Pediatric vaccine exemptions for have increased in recent years.^1^ Less is known about whether political environments may influence these rates. We studied state-level kindergarten vaccination exemption rates by 2024 Presidential election result to study changes in vaccine exemption rates from 2009-2024.

## Methods

This observational cross-sectional study was based on vaccination exemption data and state kindergarten populations obtained from the CDC.^2,3^ States (and Washington DC) were divided into Trump or Harris states based on the 2024 Presidential election results (see Supplement).^4^ Yearly incident rates (IRs) for vaccine exemptions were calculated for any exemption, medical exemptions, and non-medical exemptions with 95% confidence intervals (CIs) for the 2009-2010 through 2023-2024 seasons. Vaccination exemption incident rate ratios (IRR) were measured with 95% confidence intervals for any exemption. IRRs were considered significant if 95% CIs excluded 1.0. If states were missing data for a season, they were excluded from that season.

For each Presidential election (2012-2024)^5^, scatter plots were created to determine whether there were correlations between vaccination exemption rates and the percent of Republican vote in each election, using Spearman correlations (with corresponding p values) for non-normal data. Weighting was applied to account for state population differences.

Analyses were performed in R version 4.0.3 and Microsoft Excel 16.91. Institutional Review Board exemption was not required for these publicly available data.

## Results

The study included 53,997,748 person-years of kindergarten vaccination data. In 2023-2024, 46 (90.2%) states permitted non-medical vaccination exemptions for kindergarten children. Of states won by Donald Trump in 2024, 96.7% had non-medical exemptions (30/31). Of states won by Kamala Harris, 80% had non-medical exemptions (16/20). During the study, one Trump state (Mississippi) introduced non-medical exemptions, and four Harris states removed them.

From 2009-2024 vaccine exemptions in kindergarten children decreased in 2024 Harris states and increased in 2024 Trump states (Figure 1A). In the first season (2009-2010), children in states 2024 Harris states were more likely to have vaccination exemptions (IRR 0.93; 95% CI 0.92-0.95). Children in 2024 Trump states were more likely to have vaccine exemptions starting from the 2016-2017 season (IRR 1.30; 95% CI 1.28-1.31); IRRs increased in each subsequent season through 2023-2024 (IRR 2.53; 95% CI 2.49-2.56) (Figure 1A). Most exemptions were non-medical (Figure 1B-C).

**Figure 1.**
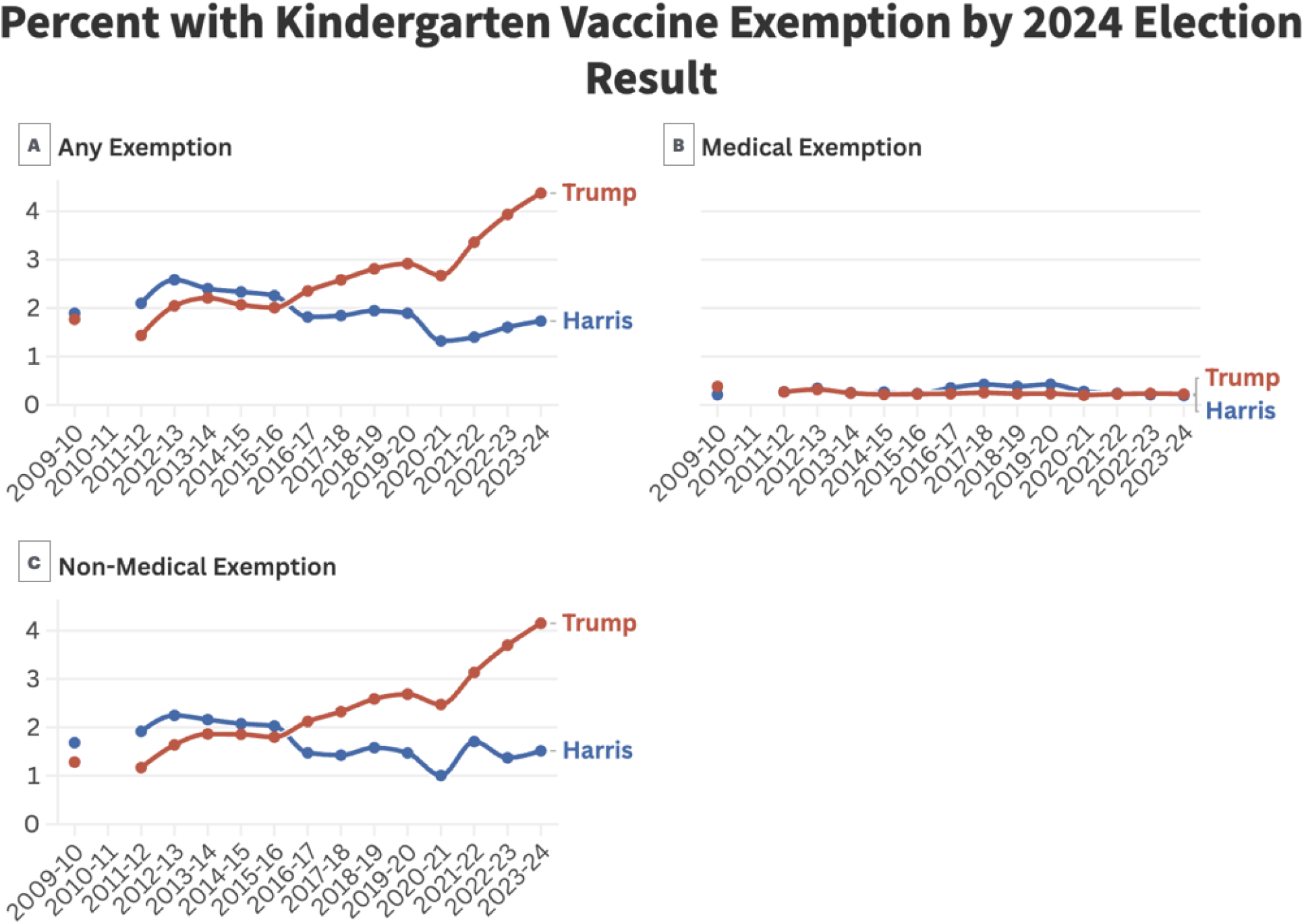
The percentage of children in kindergarten with vaccination exemptions from 2009-2010 season through the 2023-2024 season, binned by general election results in the 2024 election. Red lines correspond to states won by Donald Trump and blue lines to states won by Kamala Harris. Panel A: all exemptions; Panel B: medical exemptions only; Panel C: non-medical exemptions only. 95% confidence intervals are shown but may not be visible due to narrow bandwidths. States with missing data from certain years were excluded from both the numerator and denominators. For more information, see Supplement.

Regression coefficients comparing Republican vote in each state to vaccination exemption rates increased over the four Presidential elections assessed (Figure 2) but were not statistically significant.

**Figure 2.**
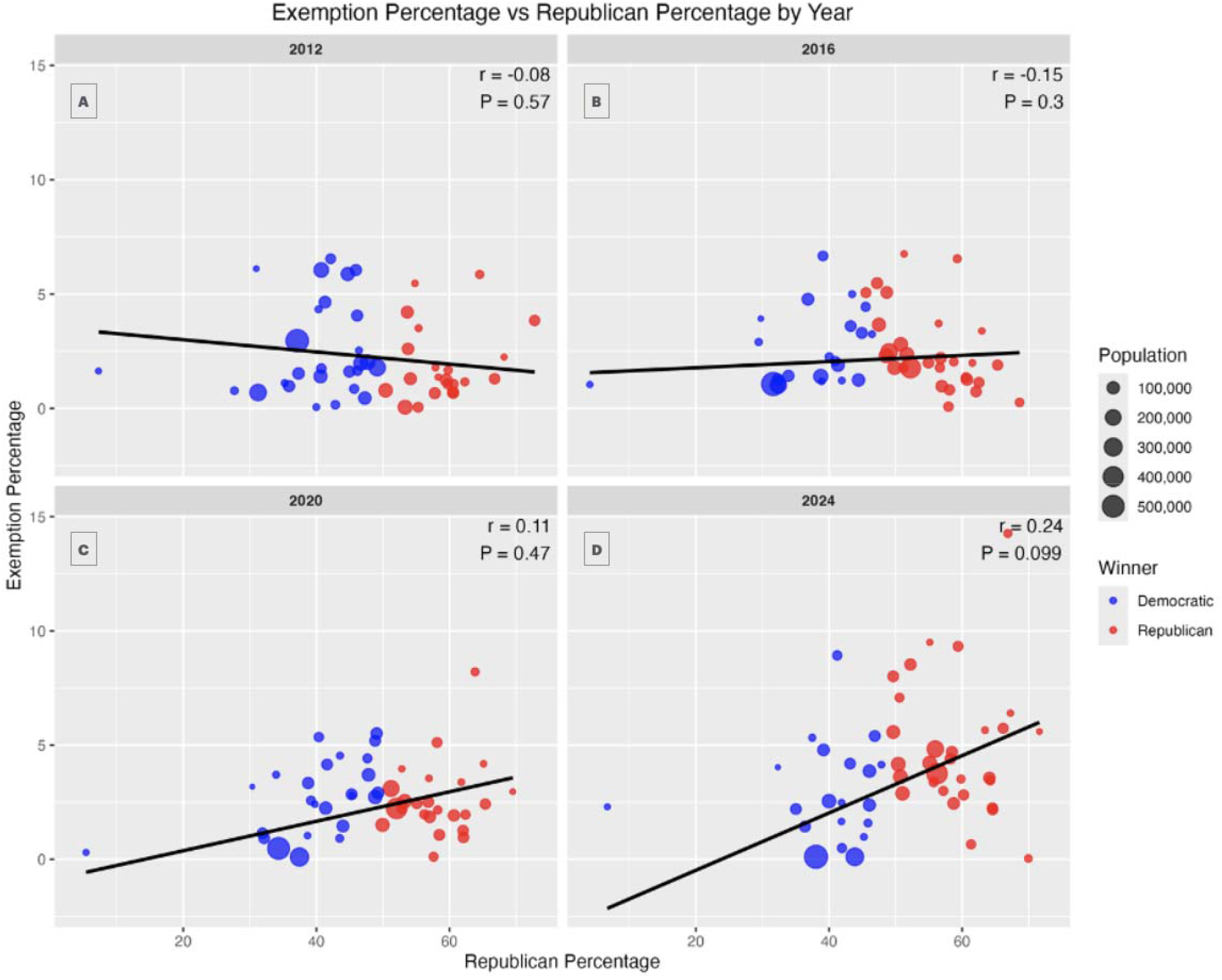
Percent of kindergarten children with vaccination exemptions versus percent of vote won by the Republican candidate in each state in four US Presidential elections. Panel A: 2012 (blue circles were states won by Barack Obama, red circles were states won by Mitt Romney); Panel B: 2016 (blue circles were states won by Hillary Clinton, red circles were states won by Donald Trump); Panel C: 2020 (blue circles were states won by Joseph Biden Obama, red circles were states won by Donald Trump); Panel D: (blue circles were states won by Kamala Harris, red circles were states won by Donald Trump). The radius of each circle corresponds to the population of children in kindergarten of each state.

## Discussion

Since 2009-2010, kindergarten vaccination exemption rates increased in eventual 2024 Trump states and decreased in Harris states, with children in Trump states >2.5-times more likely to have vaccination exemptions in 2023-2024. This change was driven by non-medical exemptions. There is no indication that medical exemptions were misused by parents or clinicians seeking to avoid vaccination in states where non-medical exemptions are banned.

An impressive change in vaccination exemption rate by political environment coincided with the 2016 election and the COVID-19 pandemic period. This may reflect increasing politicization of vaccination policies^2,3^, as mistrust of medical expertise has increased in some US regions.

Study limitations: 1) Incomplete reporting and ecological analyses; 2) Election results described establish correlation only; 3) A relatively small number of children received vaccination exemptions; it is possible that effect sizes due to political leanings may be smaller or greater; 4) In the primary analysis, 2009-2024 exemption rates were grouped by 2024 Presidential election outcome; some states won by Biden (2020), Clinton (2016), or Obama (2012) were won by Trump in 2024.

Since 2009-2010, vaccination exemption rates have increased in states won by Trump in the 2024 election, likely reflecting increasing heterogenous views stemming from the politicization of vaccination policies. This disparity appears to be new and public health efforts at local, state, and federal levels should address this alarming phenomenon.

## Data Availability

All data produced are available online at CDC.gov

## Contributions

*Concept and design:* Faust, Renton

*Acquisition, analysis, or interpretation of data:* Renton

*Drafting of the manuscript:* Faust, Renton

*Critical revision of the manuscript for important intellectual content:* Faust, Renton

*Statistical analysis:* Faust, Renton

*Obtained funding:* N.A.

*Administrative, technical, or material support:* N.A.

*Supervision:* Faust

**Supplemental materials** for: *Faust and Renton*, “Vaccination exemptions among Kindergartners by state-level 2024 Presidential election result.”

## Supplemental methods

### Grouping/binning

For each year, we summed the numerators (exemptions) and denominators (population) according to the Presidential election result. Therefore, reported percentages are raw summations for each group according to election results, not averages of the states in each group.

### CDC vaccination data caveats

- Some states do not permit nonmedical exemptions in some or all years and were not reported in the dataset. Therefore, we imputed nonmedical exemptions as 0 by subtracting medical exemptions from the number of overall (“any”) exemptions.
- There were occasional state-years in which data for one of the two categories exemption (medical or nonmedical) of were not reported in the CDC dataset, but the total number of exemptions were reported, nonetheless. In these cases, we performed simple subtraction to impute the data for the missing category.
- In one season (2021-2022), California reported 948 overall exemptions, but 1,348 medical exemptions. We assumed that this was a typographical error in the original dataset and that the number of overall exemptions was 1,348. This aligned with adjacent season trends.

